# Neurophysiological correlates of gait initiation in individuals with Huntington’s and Parkinson’s disease

**DOI:** 10.1101/2023.05.23.23290390

**Authors:** Radhika Desai, Lori Quinn

## Abstract

**Background:** Huntington’s disease (HD) and Parkinson’s disease (PD) are neurodegenerative diseases resulting in motor impairments of gait initiation (GI). However, the neurophysiological underpinnings of GI in HD and PD are not well understood. The aims of this study were to 1) evaluate the feasibility of a wireless EEG system to identify EEG-derived movement-related potentials (MRPs) in individuals with HD and PD during GI, and to 2) determine the tolerance of a high repetition GI protocol.

**New Method:** 3 participants with HD and 3 participants with PD performed 3 blocks of 15 trials of GI. EEG-derived MRPs of readiness potential (RPs) and contingent negative variation (CNV) were identified during GI, using a wireless EEG system synced with kinetic measures of GI. Tolerance of the protocol was determined from changes in COP across GI blocks.

**Results:** There were no differences between HD and PD for CNV and RP amplitudes and latencies, although they were within acceptable ranges of MRP values in HD and PD. There were no differences of COP values between GI blocks.

**Comparison with old method:** A wireless EEG system elicits naturalistic GI biomechanical responses as opposed to previous methods employing tethered systems. In contrast to single-trial EEG studies, this study implemented a larger number of GI trials, which produces greater MRP resolution.

**Conclusions:** This study validated the use of a wireless EEG headset in determining MRP values during GI in HD and PD. Participants within these populations were able to tolerate a high repetition GI protocol. Future work will explore MRPs in larger cross-sectional studies for the development of clinical outcome measures.

## Introduction

Huntington’s disease (HD) and Parkinson’s disease (PD) are neurodegenerative diseases causing dysfunction and death of cells within the basal ganglia, and thus disruption of pathways with resultant impairment of cognition, motor function, and behavior (1,2). Among the many motor deficits in HD and PD, presentation of impaired movement initiation, and specifically gait initiation, is a hallmark characteristic of both diseases (3,4).

Gait initiation is a complex motor task involving volitional and non-volitional components. It is comprised of anticipatory postural adjustments (APAs) through first step completion (5,6). APAs precede first step execution and consist of two mediolateral components involved with loading and unloading of the stance leg, and an anteroposterior component for backwards weight shift and acceleration into the first step. APAs are the non-volitional, or implicit, component of gait initiation needed to maintain postural and trunk stability as an individual initiates and executes the first step(7). APAs in non-neurological populations are scaled to match the force and acceleration demands of gait initiation. In HD and PD populations, APAs exhibit accelerations, durations, and displacements that are too large or too small for movement demands (8). The impaired ability to modulate spatial temporal parameters of components of gait initiation is rooted in the inability to appropriately modulate force (3,9). The neuropathology behind force modulation errors, specifically in basal ganglia disorders, are not well understood. Understanding the neural underpinnings of force modulation errors can provide greater insight into the role of the basal ganglia and its contribution to gait impairments in people with HD and PD.

Electroencephalography (EEG) is a neurophysiological measurement method that records neuronal oscillations in real-time. The high temporal resolution of this method allows for recording of brain activity during or prior to a movement. Specifically, movement-related potentials (MRPs) are time-locked event-related potentials that occur just prior to movement onset (10). Specifically, contingent negative variation (CNV) and the readiness potential (RP) are two MRPs reflective of two stages of motor planning. Contingent negative variation is a MRP that is a measure of attention allocation to movement intent and is a far field-potential of activity in pre-SMA areas. The RP follows the CNV and is reflective of motor planning just prior to motor execution. The RP is measured from the SMA and is generated by basal-ganglia-thalamocortical drive (10). Measurements of MRPs provide insight into the temporal impairments of the basal ganglia and motor planning areas of the brain in individuals with HD and PD.

Previous methods of MRP measurements during task execution involved the use of a tethered or wired system. These systems consisted of setups in which a participant wore a headset with wires that reached an external amplifier system or wore the amplifier and receiver in a backpack-like design. Although wired systems can provide a greater sampling rate, these systems often offset an individual’s center of mass in the posterior direction, thereby limiting naturalistic and valid motor task execution. ecent advancements in mobile EEG devices enable evaluation of neural patterns during gait initiation without disrupting an individual’s natural movement pattern. These wireless EEG systems are lightweight and transmit signal via Bluetooth and an external sync unit, maintaining a participant’s normal biomechanical characteristics. Previous research has attempted to identify distinct EEG patterns in people with early-stage HD and PD, however these study designs had limitations in their ability to adequately elicit motor planning responses, and naturalistic motor execution (11,12). Gait initiation is a complex motor task that places greater demands on motor planning areas on the brain, relative to simple upper extremity tasks.

Specifically, the supplementary motor area (SMA) is a primary cortical area of motor planning involved with temporal and spatial scaling of motor tasks. Gait initiation involves a high temporal and spatial scaling acuity, thereby placing a greater demand on SMA processing. Therefore, gait initiation likely elicits more measurable responses from the SMA as MRPs. Furthermore, early changes in gait initiation parameters among HD and PD are likely to elicit a sensitive MRP measure of basal-ganglia-thalamocortical (BG-SMA) dysfunction. However, the feasibility and tolerance to high repetition of gait initiation trials needed for reasonable resolution of MRPs has yet to be assessed in HD and PD populations. This approach of linking motor behavior to real-time neural signal will also elucidate the role of the basal ganglia in motor planning of gait initiation, which has mechanistic implications for both neurological and non-neurological motor control processes.

The aims of this study were to: 1) to determine the feasibility of using a wireless EEG system to identify EEG-movement related potentials of individuals of HD and PD during gait initiation and to 2) determine the tolerance of a high repetition gait initiation protocol. Ultimately this study will enable us to evaluate more complex motor tasks with a m more feasible protocol,

## Materials and Methods

### Participants

For this pilot study, 3 individuals with manifest HD, 3 individuals with mid stage PD participated in this study. The following methodology covers methods used to collect data on these six participants but is also the intended methodology for future data collections. HD participants were recruited through an existing registry established by the Neurorehabilitation Research Laboratory at Teachers College Columbia University and through the Huntington’s Disease Society of America Center of Excellence at Columbia University and the PD Registry at Teachers College. HD and PD participants had disease status confirmed by a neurologist. A sample size of 15 HD and 15 PD was determined by modeling after previous studies examining movement related potentials in HD and PD participants during motor execution tasks (13,14). However, due to Covid-19 lockdowns, only pilot data was collected for 3 HD participants and 3 PD participants. All 3 PD participants were on a medication-on status (Levodopa).

### Inclusion and exclusion criteria

The inclusion and exclusion criteria of Huntington’s participants were 1) age 18 years of age or older 2) genetically confirmed HD, 3) a total Functional Capacity Score ≥7, and the ability to walk 2 blocks without an assistive device. For PD participants the inclusion and exclusion criteria are 1) Age 18 years of age or older, 2) clinical diagnosis of PD, 3) Hoehn & Yahr stage I or II 4) able to walk 2 blocks without an assistive device. Participant demographics are listed in table 1.1.

**Table 1.1:**
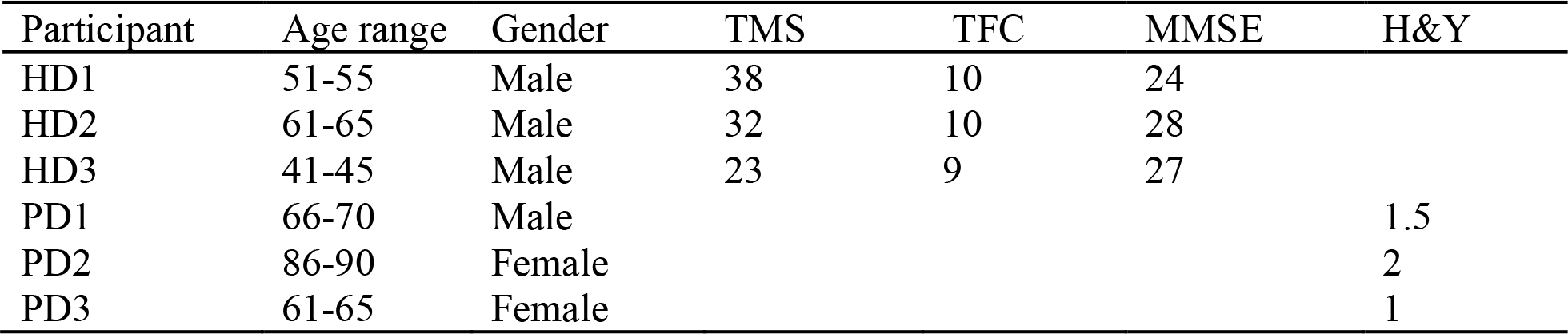
Participant demographics Table 1.1 Displays age, gender, total motor scores (TMS), total functional capacity (TFC), and Mini Mental State Examination (MMSE) for each HD and PD participant. Unified Huntington’s Disease Rating Scale (UHDRS) total motor score (TMS) is a sum of all sub scores rated on a 0-4 scale, where 0 represents an absence of impairment and 4 represents a pronounced impairment. UHDRS Functional Assessment (FA) scores are out of 25, where larger values represent higher functioning, and independence (Ind) scores, where higher scores represent greater independence, are rated 0-100. The MMSE is a 30 point test that evaluates cognitive functioning including attention, orientation, memory and language. Hoehn and Yahr (H&Y) scores for PD participants are listed in the final column, where a score of 2 or below represents an early stage of the disease.

#### Materials and Setup

A 32-channel wireless scalp EEG system (B-Alert, Advanced Brain Monitoring, Carlsbad, CA) were used to record brain activity throughout each trial. The electrode array of the EEG system is embedded within a plastic sensor strip. Preparation of the array involves placing foam attachments to each channel site for the purposes of holding salt solution. A salt and water conductance gel were placed along each sponge site on each electrode prior to placement on the participant. Reference electrodes were adhered to participants mastoid processes bilaterally and clipped into place to the sensor strip. Velcro straps were attached to flexible bands, which hold the sensor strips and are adjustable to participant head size. The sensor strip was placed on each participant using the 10-20 system. Impedance was checked through the B-Alert software system and the following thresholds was used to assess thresholds: Impedance values < 40kΩ were highlighted in green, values 40kΩ -80kΩ will be yellow, and impedance values > 80kΩ were red (indicating that the sensor is outside the acceptable range). The external receiver for the EEG headset was calibrated to establish connection with the external sync unit (ESU), which receives the signal via Bluetooth and is connected to the main computer dock via USB. Calibration of the ESU was performed within the B-Alert software.

An embedded Kistler force plate (Kistler Instrument Corp, Novi, MI) was used to measure kinetics of gait initiation trials. The force plate was calibrated and centered using dimension adjustments within the Vicon Nexus software system with no additional gain. Furthermore, the force plate was reset to zero within Vicon and at the level of the hardware from the amplifier. Sampling was synchronized with a hardware sync using the ESU and force place amplifier, allowing EEG data to be time-locked with kinetic markers of gait initiation.

### Protocol

Each participant performed 60 self-initiated walking trials. Participants began standing with both feet on an embedded Kistler force plate. They were instructed to begin walking when ready, and walk until they reached a mark placed on the floor 5-m in front of their starting position. No further cues were provided as we aim to understand internally generated gait initiation. Participants were instructed to walk 5 m because MRPs for forward stepping are different from the initiation of continues gait (Figure 3).

### Data Processing

EEG data from each trial was filtered and processed using Matlab (Matlab, Mathworks, Natick, MA). Filter settings and processing are modeled from previous studies that determined MRPs from gait initiation. All EEG trials were resampled to fit the 1000hz sampling frequency of the synced force plate. A zero-phase bandpass filter with a range of 8-40hz was applied to each raw EEG trial for each subject. We used EEG the CZ channel electrode for MRP extraction consistent with previous literature measuring MRP during gait initiation (13–15).

MRPs were found using the epochs established and indices found from APA events within the kinetic gait initiation data. The CNV occurs within the 400 ms window prior to initiation, and the RP occurs within the 200 ms window prior to gait initiation. These time thresholds are based on epochs that were previously used in an HD population when identifying the CNV and RP. Specifically, Turner et. al, 2015 used these same time thresholds prior to movement onset in pre-manifest and manifest-HD participants. This window was also chosen as it is most associated with the initiation of movement rather than processing related to cue onset. It was important to create MRP windows most associated with motor planning of movement initiation rather than the sensorimotor planning stage that follows cue onset. MRPs built around cue onset would have implications for PD participants as changes in neurophysiology and behavior are influenced by cued vs uncued paradigms. CNV and RPs were identified using the start of the first mediolateral APA for one set of MRPs, and the start of heel-off as the second set of MRPs. Using both the first APA and heel-off as the end index for MRPs will provide insight on the motor planning associated with the APA and heel-off associated with the first step.

Additionally, methodologies in previous studies identify MRPs during gait initiation have used both APA start and heel-off as end index markers for MRPs (16). Peak amplitudes and latencies, as onset times of peak amplitudes relative to gait initiation start, of the CNV and RP served as outcome variables.

CNV amplitudes were extracted as the maximum value (microvolts **μ**V) within the CNV time window prior to the onset of the first mediolateral APA and heel-off, while latencies were defined as the time of onset (milliseconds ms) of the CNV amplitude relative to both movement onsets. All mean amplitudes and latencies for RPs and CNVs prior to APA and heel-off are listed in table 2 and table 3 respectively. RP amplitudes and latencies were determined similarly to CNV values, with the time window following CNV offset. Mean values, standard deviations, mean differences between groups with confidence intervals, and effect sizes were determined for amplitudes and latencies for HD and PD participants. For CNV amplitudes, HD (M = 13.87, SD = 6.82) and PD (M = 13.95, SD = 6.35) exhibited similar mean values with a very small effect size (d=0.01). Consistently, CNV latencies prior to the APA were also similar between HD (M=141.54, SD=24.15) and PD (M=141.85, SD=24.36, d=0.01).

**Table 1.2:** Individual data for MRPs prior to APA onset Table 1.2: Contingent negative variation (CNV) amplitudes and latencies along with readiness potential (RP) amplitudes and latencies are listed for each participant. Each value represents the mean of value across all trials within the participant. Amplitudes are maximum values in microvolts from the Cz channel prior to the onset of the first APA. Latencies are the onset of each amplitude relative to APA onset.

| Participant | CNV amplitude($\mu V$) | CNV latency (ms) | RP amplitude ($\mu V$) | RP latency (ms) |
| --- | --- | --- | --- | --- |
| HD1 | 14.09(6.83) | 142.58(23.71) | 12.51(7.02) | 39.89(9.09) |
| HD2 | 14.33(9.73) | 130.33(20.76) | 10.03(6.41) | 40.20(10.32) |
| HD3 | 13.10(4.91) | 144.37(25.68) | 9.57(4.88) | 38.87(8.57) |
| PD1 | 12.11(6.58) | 134.95(23.60) | 9.59(5.11) | 38.23(10.23) |
| PD2 | 14.64(5.98) | 140.39(25.63) | 11.36(6.48) | 40.04(9.00) |
| PD3 | 13.47(6.58) | 142.84(23.57) | 11.20(6.27) | 41.45(9.32) |

**Table 1.3:** Group data for MRPs prior to APA Table 1.3: Mean values for contingent negative variation (CNV) amplitudes and latencies, and readiness potential (RP) amplitudes and latencies prior to APA onset are listed for PD and HD groups. Mean differences between groups are listed along with their confidence intervals (CI) at 95%. Effect sizes are listed for mean differences represented by Cohen’s D.

|  | PD mean (SD) | HD mean (SD) | Mean Differences (CI) | Effect size (Cohen's D) |
| --- | --- | --- | --- | --- |
| CNV amplitude ($\mu V$) | 13.75(6.22) | 13.87(6.82) | 0.12(-1.67-1.41) | 0.02 |
| CNV latency (ms) | 140.85(24.00) | 141.54(24.15) | 0.69(-6.32-4.96) | 0.03 |
| RP amplitude ($\mu V$) | 11.06(5.96) | 11.51(6.62) | 0.45(-1.93-1.04) | 0.07 |
| RP latency (ms) | 40.48(9.40) | 39.68(9.10) | -0.8(-1.36-2.96) | 0.09 |

**Table 1.4:**
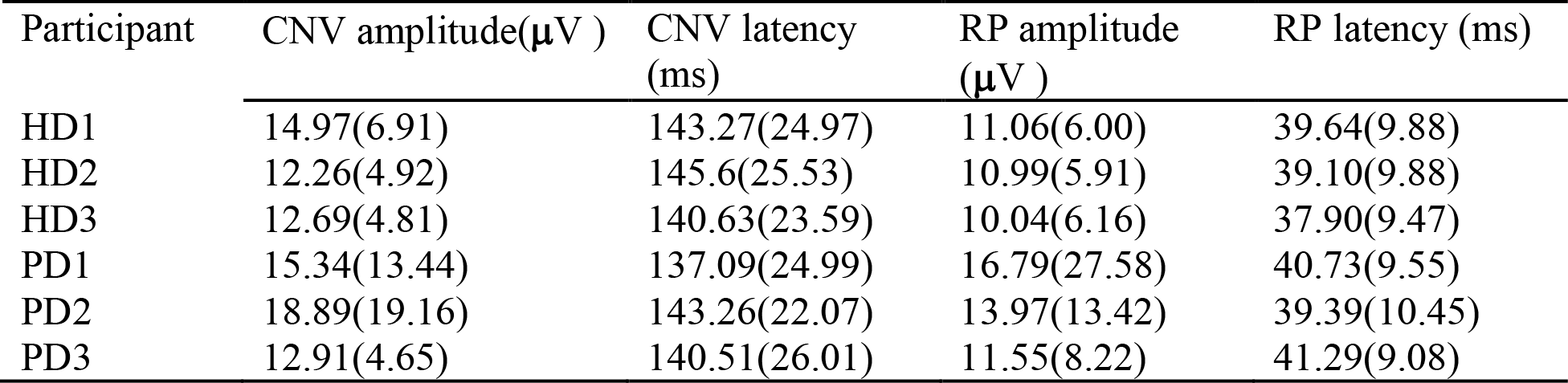
Individual data for MRPs prior to heel-off Table 1.4: Contingent negative variation (CNV) amplitudes and latencies along with readiness potential (RP) amplitudes and latencies are listed for each participant. Each value represents the mean of value across all trials within the participant. Amplitudes are maximum values in microvolts from the Cz channel prior to the onset of heel-off of the stepping leg. Latencies are the onset of each amplitude relative to APA onset.

| Participant | CNV amplitude($\mu V$) | CNV latency (ms) | RP amplitude ($\mu V$) | RP latency (ms) |
| --- | --- | --- | --- | --- |
| HD1 | 14.97(6.91) | 143.27(24.97) | 11.06(6.00) | 39.64(9.88) |
| HD2 | 12.26(4.92) | 145.6(25.53) | 10.99(5.91) | 39.10(9.88) |
| HD3 | 12.69(4.81) | 140.63(23.59) | 10.04(6.16) | 37.90(9.47) |
| PD1 | 15.34(13.44) | 137.09(24.99) | 16.79(27.58) | 40.73(9.55) |
| PD2 | 18.89(19.16) | 143.26(22.07) | 13.97(13.42) | 39.39(10.45) |
| PD3 | 12.91(4.65) | 140.51(26.01) | 11.55(8.22) | 41.29(9.08) |

**Table 1.5:** Group data for MRPs prior to heel-off Table 1.5: Mean values for contingent negative variation (CNV) amplitudes and latencies, and readiness potential (RP) amplitudes and latencies prior to heel-off are listed for PD and HD groups. Mean differences between groups are listed along with their confidence intervals (CI) at 95%. Effect sizes are listed for mean differences represented by Cohen’s D.

|  | PD mean (SD) | HD mean (SD) | Mean differences (CI) | Effect size (Cohen's D) |
| --- | --- | --- | --- | --- |
| CNV amplitude ($\mu\text{V}$) | 15.42(13.30) | 13.96(6.26) | -1.46(-1.23-4.14) | 0.15 |
| CNV latency (ms) | 141.12(24.36) | 143.21(24.70) | 2.09(-8.14-3.94) | 0.09 |
| RP amplitude ($\mu\text{V}$) | 13.40(15.51) | 10.84(5.99) | -2.56(-0.50-5.62) | 0.23 |
| RP latency (ms) | 40.65(9.61) | 39.18(9.75) | -1.47(-0.91-3.85) | 0.15 |

### Statistical analysis

For the pilot data collected, descriptive statistics (mean, SD) were calculated for all COP measures, CNV amplitudes and latencies, and RP amplitudes and latencies. Mean differences with effect sizes and 95% confidence intervals (CI) were also calculated for differences between HD and PD participants across all variables. Effect sizes were calculated using Cohen’s D, where moderate effect sizes were indicated by values above .50, and large effect sizes were indicated by values above .80. Values below .50 suggested no difference between means. The initial statistical analysis plan involved determining group differences for CNV amplitudes, CNV latencies, RP amplitudes, and RP latencies using a one-way ANOVA between HD, PD, and non-neurological controls.

## Results

All mean amplitudes and latencies for RPs and CNVs prior to APA and heel-off are listed in table 1.2 and table 1.3 respectively. RP amplitudes and latencies were determined similarly to CNV values, with the time window following CNV offset. Mean values, standard deviations, mean differences between groups with confidence intervals, and effect sizes were determined for amplitudes and latencies for HD and PD participants (Table 1.2). For CNV amplitudes, HD (M = 13.87, SD = 6.82) and PD (M = 13.95, SD = 6.35) exhibited similar mean values with a very small effect size (d=0.01). Consistently, CNV latencies prior to the APA were also similar between HD (M=141.54, SD=24.15) and PD (M=141.85, SD=24.36, d=0.01).

### MRPs for prior to heel-off

For CNV outcome measures prior to heel-off PD exhibited slightly large amplitudes (M = 15.42, SD=13.30, d=0.15) than HD (M=13.96, SD=6.26). PD also displayed slightly smaller latency values (M=141.12, SD=24.36) relative to HD (M=143.21, SD=24.70) with very small effect size (d=0.09). RP amplitudes prior to APA onset were similar between PD (M= 11.25, SD=6.33) and HD (M=11.51, SD=6.62, d= 0.04). However, for RP latencies prior to APA onset, PD exhibited slightly earlier latencies (M=40.88, SD=9.18), while HD exhibited slightly later latencies (M=39.68, SD-9.10, d=0.13). RP amplitudes prior to heel-off indicated larger values among PD participants (M=13.40, SD=15.51) than HD participants (M=10.84, SD=5.99) with a moderate effect size. Similar to RP latencies prior to the APA, PD exhibited earlier RP latencies prior to heel-off (M=40.65, SD= 9.61) as compared to HD participants (M=39.18, SD=9.75, d=0.15) with no difference.

## Conclusion

The purpose of this study was to determine the feasibility of a wireless EEG system in identifying real-time movement related potentials of gait initiation in individuals with HD and PD, and tolerance to a high repetition gait initiation protocol in these populations. Movement-related potentials revealed temporal characteristics of motor planning and elucidated inhibitory and excitatory dynamics that precede motor execution. The MRP values obtained from the wireless EEG system were within acceptable ranges as compared to previously determined wired or tethered EEG systems. We used a portable EEG system, which was time locked with an embedded force plate to record both neural and kinetic in real-time among three individuals with mid-stage PD and three individuals with manifest HD. Epochs prior to gait initiation for EEG and kinetic data were built using software annotation markers associated with the cue provided to participants, and kinetic markers of the APA and heel-off during post processing. CNV and RPs were the movement-related potentials of interest that were extracted during post processing and reflect frontal lobe projections and basal-ganglia-thalamo-cortical projections respectively.

Results confirmed previous findings for kinetic parameters and validated the methodology in its ability to measure movement-related potentials prior to gait initiation in HD and PD populations. No study to date has used wireless EEG technology to measure neural signal in real-time during gait initiation in PD and HD. However, the values obtained from this system and methods were similar to the results determined in wired and tethered systems.

Most knowledge regarding the state of basal-ganglia-thalamo cortical function in PD and HD are largely from static imaging studies (17–19). Although brain morphology during static conditions can provide us with an understanding of how PD and HD progression affect neuroanatomy, static imaging alone cannot unveil the changes that occur in structural dynamics. Compensatory mechanisms of neuroplasticity may prevent isolated degradation of neuroanatomy from presenting both functionally and neurophysiologically (20). Therefore, measuring temporal dynamics of neuronal activity has the capacity to reveal changes in brain functioning that may not otherwise be captured in imaging techniques. Specifically, neural patterns related to motor planning are most appropriate for measurements with high temporal resolution, as excitatory regulation, and timing of this is critical for successful movement onset (21,22). EEG has the ability to measure real-time recordings of neuronal activity prior to movement thereby capturing true movement planning as revealed by brain dynamics. The strength of our study was that it used EEG during gait initiation, a complex motor task that places large demands on motor planning and motor cortical systems. Unlike simple upper extremity tasks used in previous studies to measure MRPs, gait initiation involves coordination of whole-body dynamics, and appropriate postural responses to initiate displacement of the body. Gait initiation also involves many of the brain structures most vulnerable to PD and HD progression, thereby making it an optimal task in eliciting measurable motor planning responses.

We used 4 blocks of 15 gait initiation trials per subject to optimize the temporal resolution of the EEG recordings without taxing participants. These 60 trials allowed for an increase in MRP resolution and a decrease of noise, as a greater amount of trials are averaged to generate composite amplitudes and latencies (23). We found that participants were able to tolerate 60 trials of gait initiation, when trials were comprised of blocks, and that this trial number adequately generated CNV and RP responses. Tolerance to the task and EEG system by both PD and HD participants was also due to the EEG system we used. The B-Alert monitoring system is a lightweight wireless EEG system that is secured on a participant’s scalp via Velcro straps. We believe the noninvasive and light nature of this EEG system minimizes the possibility of patient discomfort. Additionally, this untethered and lightweight EEG system facilitated natural and free walking for participants, unlike many tethered EEG systems that limit a participant into moving a single or several steps. The ability to freely walk with this protocol was necessary as MRPs associated with walking are separate from that of MRPs associated with step execution. We further capitalized on this notion by instructing participants to walk approximately 10 meters with ongoing EEG recording, with no cue or disturbance provided after first step completion. The combination of the EEG system and cues provided to participants allowed for a valid recording of neural signals prior to the planning and execution of true gait.

MRPs have the capacity to be a feasible and cost-effective tool that reveals changes in neural circuitry dynamics thereby also providing information regarding neural integrity. Noninvasive biomarkers, such as MRPs, would also allow clinicians to more easily identify pre-HD and early-stage PD individuals that are closer to the manifestation of symptoms, ultimately optimizing clinical management. An ideal biomarker for disease is one that is sensitive to, and highly correlated with disease progression and respective pathophysiology. Therefore, the task of determining neurophysiological mechanisms underlying gait impairments in HD remains critical.

## Data Availability

All data produced in the present study are available upon reasonable request to the author.

## Notes

### Competing Interest Statement

The authors have declared no competing interest.

### Funding Statement

This study did not receive any funding.

### Author Declarations

This research protocol was approved by the Institutional Review Boards at Teachers College Columbia University and through the Huntingtons Disease Society of America Center of Excellence at Columbia University and the PD Registry at Teachers College.

## References

1. Roos RA. Huntington’s disease: a clinical review. Orphanet J Rare Dis. 2010 Dec;5(1):40.

2. Underwood CF, Parr-Brownlie LC. Primary motor cortex in Parkinson’s disease: Functional changes and opportunities for neurostimulation. Neurobiology of Disease. 2021 Jan;147:105159.

3. Delval A, Krystkowiak P, Blatt JL, Labyt E, Bourriez JL, Dujardin K, et al. A biomechanical study of gait initiation in Huntington’s disease. Gait and Posture. 2007;25(2):279–88.

4. Hass CJ, Waddell DE, Fleming RP, Juncos JL, Gregor RJ. Gait Initiation and Dynamic Balance Control in Parkinson’s Disease. Archives of Physical Medicine and Rehabilitation. 2005 Nov;86(11):2172–6.

5. Bloedel JR, Ebner TJ, Wise SP, editors. The acquisition of motor behavior in vertebrates. Cambridge, Mass: MIT Press; 1996. 440 p.

6. Delval A, Krystkowiak P, Blatt JL, Labyt E, Destée A, Derambure P, et al. Differences in anticipatory postural adjustments between self-generated and triggered gait initiation in 20 healthy subjects]. Neurophysiologie clinique = Clinical neurophysiology. 2006;35(5– 6):180–90.

7. MacKinnon CD, Bissig D, Chiusano J, Miller E, Rudnick L, Jager C, et al. Preparation of anticipatory postural adjustments prior to stepping. Journal of neurophysiology. 2007;97(6):4368–79.

8. Roemmich RT, Nocera JR, Vallabhajosula S, Amano S, Naugle KM, Stegemöller EL, et al. Spatiotemporal variability during gait initiation in Parkinson’s disease. Gait & Posture. 2012 Jul;36(3):340–3.

9. Desai R, Fritz NE, Muratori L, Hausdorff JM, Busse M, Quinn L. Evaluation of gait initiation parameters using inertial sensors in Huntington’s Disease: Insights into anticipatory postural adjustments and cognitive interference [Internet]. Rehabilitation Medicine and Physical Therapy; 2020 Aug [cited 2021 Apr 8]. Available from: http://medrxiv.org/lookup/doi/10.1101/2020.08.13.20174235

10. Jahanshahi M, Hallett M, editors. The Bereitschaftspotential [Internet]. Boston, MA: Springer US; 2003 [cited 2021 Jun 17]. Available from: http://link.springer.com/10.1007/978-1-4615-0189-3

11. Nguyen L, Bradshaw JL, Stout JC, Croft RJ, Georgiou-Karistianis N. Electrophysiological measures as potential biomarkers in Huntington’s disease: Review and future directions. Vol. 64, Brain Research Reviews. 2010. p. 177–94.

12. Turner LM, Croft RJ, Churchyard A, Looi JCL, Apthorp D, Georgiou-Karistianis N. Abnormal electrophysiological motor responses in Huntington’s disease: Evidence of premanifest compensation. PLoS ONE. 2015;10(9).

13. Shoushtarian M, Murphy A, Iansek R. Examination of central gait control mechanisms in Parkinson’s disease using movement-related potentials: MRP Disturbances in Parkinson Gait. Mov Disord. 2011 Nov;26(13):2347–53.

14. Turner LM, Croft RJ, Churchyard A, Looi JCL, Apthorp D, Georgiou-Karistianis N. Abnormal Electrophysiological Motor Responses in Huntington’s Disease: Evidence of Premanifest Compensation. Gonzalez-Alegre P, editor. PLoS ONE. 2015 Sep 25;10(9):e0138563.

15. Rektor I, Bareš M, Kubová D. Movement-related potentials in the basal ganglia: A SEEG readiness potential study. Clinical Neurophysiology. 2001;

16. Nascimento OFD, Nielsen KD, Voigt M. Influence of directional orientations during gait initiation and stepping on movement-related cortical potentials. Behavioural Brain Research. 2005;

17. Filippi MM, Oliveri M, Pasqualetti P, Cicinelli P, Traversa R, Vernieri F, et al. Effects of motor imagery on motor cortical output topography in Parkinson’s disease. Neurology. 2001 Jul 10;57(1):55–61.

18. Guo Q, Wang D, He X, Feng Q, Lin R, Xu F, et al. Whole-Brain Mapping of Inputs to Projection Neurons and Cholinergic Interneurons in the Dorsal Striatum. Arenkiel B, editor. PLoS ONE. 2015 Apr 1;10(4):e0123381.

19. Zeun P, Scahill RI, Tabrizi SJ, Wild EJ. Fluid and imaging biomarkers for Huntington’s disease. Mol Cell Neurosci. 2019 Jun;97:67–80.

20. Klöppel S, Gregory S, Scheller E, Minkova L, Razi A, Durr A, et al. Compensation in Preclinical Huntington’s Disease: Evidence From the Track-On HD Study. EBioMedicine. 2015 Oct;2(10):1420–9.

21. Schippling S, Schneider SA, Bhatia KP, Münchau A, Rothwell JC, Tabrizi SJ, et al. Abnormal Motor Cortex Excitability in Preclinical and Very Early Huntington’s Disease. Biological Psychiatry. 2009 Jun;65(11):959–65.

22. Shakeel A, Navid MS, Anwar MN, Mazhar S, Jochumsen M, Niazi IK. A Review of Techniques for Detection of Movement Intention Using Movement-Related Cortical Potentials. Computational and Mathematical Methods in Medicine. 2015;2015:1–13.

23. Scheer HJ, Sander T, Trahms L. The influence of amplifier, interface and biological noise on signal quality in high-resolution EEG recordings. Physiol Meas. 2006 Feb 1;27(2):109–17.

